# Decentralized governance may lead to higher infection levels and sub-optimal releases of quarantines amid the COVID-19 pandemic

**DOI:** 10.1101/2020.05.20.20108167

**Authors:** Adam Lampert

**Affiliations:** Institute of Environmental Sciences, The Robert H. Smith Faculty of Agriculture, Food and Environment, The Hebrew University of Jerusalem, Rehovot, Israel

**Keywords:** COVID-19, dynamic games, international policy, quarantine policy

## Abstract

The outbreak of the novel Coronavirus (COVID-19) has led countries worldwide to administer quarantine policies. However, each country or state decides independently what mobility restrictions to administer within its borders, while aiming to maximize its own citizens’ welfare. Since individuals travel between countries and states, the policy in one country affects the infection levels in other countries. Therefore, major question is whether the policies dictated by multiple governments could be efficient. Here we focus on the decision regarding the timing of releasing quarantines, which were common during the first year of the pandemic. We consider a game-theoretical epidemiological model in which each government decides when to switch from a restrictive to a non-restrictive quarantine and vice versa. We show that, if travel between countries is frequent, then the policy dictated by multiple governments is sub-optimal. But if international travel is restricted, then the policy may become optimal.

## Introduction

The outbreak of the novel Coronavirus (COVID-19) necessitated quarantine policies, particularly during its early stages, before vaccines were available [1-4]. While mobility restriction measures have been taken during several disease outbreaks in history [5-11], the quarantines needed for the COVID-19 pandemic were at global scales that encompass entire countries. Such quarantines have economic and social costs [12, 13], and major relevant questions are how restrictive the quarantines should be, and what would be the right timing to release some of the mobility restrictions [1-3]. In practice, this decision is made independently by multiple countries [14-16], or independently by multiple states in countries like the U.S., or even independently by multiple municipal authorities [17]. Each governor might incline to dictate the strategy that best serves her/his own citizens; however, in periods when the quarantines are less restrictive, travelers can transmit the disease between countries, states, and cities. Consequently, the quarantine policy in one country/state may ultimately affect the outcome in other countries/states.

Such decentralized governance has a benefit: Each country or state may have a better knowledge of its own citizens’ lifestyle and needs and may dictate a policy that better suits its own citizens. However, the decentralized policy also comes with a cost: Each government might ignore the cost borne to citizens of other countries due to international and interstate travel. Accordingly, various previous game theory studies have suggested that agents (individuals or countries) under-invest in the prevention and control of diseases [18-23].

In this paper, we examine the case where each governor decides independently about the timing of releasing the quarantine, and we ask what the inefficiency is due to such decentralized governance. Namely, we examine how the strategies of different governments differ from the socially optimal strategy of a hypothetical centralized government that aims to maximize the welfare of all the citizens in all the countries. Specifically, we consider two countries/states, and we analyze the following three cases (Fig. 1) [14-16, 24]: (1) the countries have approximately the same population size and infection level; (2) the countries have approximately the same population size, but one country experiences a more severe outbreak at a given point in time; and (Case 3) one state experiences a more severe outbreak compared to the rest of the country.

**Figure 1:**
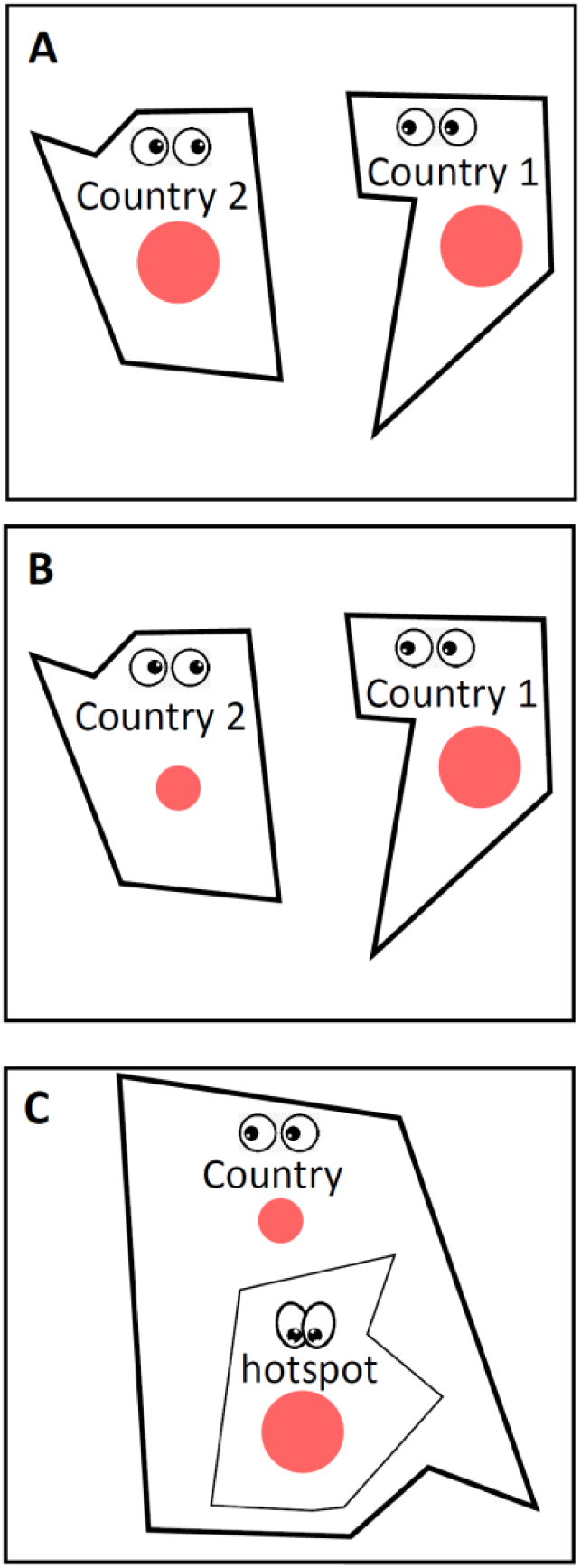
Illustration of the three cases that we consider. (A) Case 1: Both countries have the same population size and infection level. (B) Case 2: Both countries have the same population size, but country 1 has a more severe outbreak. (C) Case 3: One U.S. state is a “hotspot” and has a more severe outbreak than the rest of the U.S.

## Model

Our model is general, but some of the assumptions and parameterization are motivated by the COVID-19 outbreak. We consider two states or countries, 1 and 2, and in line with data about early COPVID-19 outbreaks [4, 14, 16, 24-27], we assume that the number of infected people in each country is very small compared to the country’s total population size. Accordingly, we consider only the dynamics of the infection level in each country, *I*_*i*_, defined as the portion of individuals that are infected in country *i*. (Namely, in contrast to traditional SIR models [28], here we consider shorter timescales during which the number of susceptible individuals is constant, which is in line with the COVID-19 data from 2020, when quarantines were common [4, 14, 16, 24-27]).

We assume for simplicity that each government can administer one of two types of quarantine at any given time: restrictive and non-restrictive. Each government can choose when to switch between these two quarantines. We assume that under a restrictive quarantine in country *i, I*_*i*_ decreases exponentially after the first two weeks at a rate −*r*_0_ as long as *I*_*i*_ > *I*_min_. (Even a restrictive quarantine is not expected to eliminate the disease entirely, and *I*_min_ characterizes some minimal infection level that persists in the population.) In turn, according to evidence showing that the infection level may still increase under non-restrictive quarantine conditions [4, 14, 16, 26, 27], we assume that *I*_*i*_ increases at a rate *r*_*i*_ under a non-restrictive quarantine, where *r*_*i*_ is country-specific and can be higher in those countries or states where interactions among individuals are more frequent. Also, we assume that if a country is under a restrictive quarantine, there is no travel from or to that country, whereas if both countries are under a non-restrictive quarantine, some individuals travel between these countries. We describe in more detail the dynamics of *I*_1_ and *I*_2_ in Methods: Dynamics of the infection levels.

In turn, the government in each country dictates the timing at which it switches from the non-restrictive quarantine to the restrictive one and vice versa. We assume that a government will not allow the health system in its country to collapse [29], and therefore, it will always switch to a restrictive quarantine if *I*_*i*_ approaches some maximum capacity, *I*_*i*_ = *I*_max_. In turn, under this constraint, the objective of each government is to maximize the proportion of time during which its own citizens are under a non-restrictive quarantine. We describe in more detail the objective functions in Methods: Objective functions.

Finally, we calculate the strategies of the governments in a Nash equilibrium and compare them to the socially optimal solution. In general, the Nash equilibria in such a dynamic game depend on various assumptions about the information that each government has [30]. Here we assume that each government does not get feedback about the infection level in the other country/state and decides in advance when it releases the quarantine. In our model, this results in a unique Nash equilibrium per a given set of parameter values (open-loop Nash equilibrium [30]). The numerical method that we used for finding the open-loop Nash equilibria and the optimal solutions is described in Methods: Numerical methods.

## Results & Discussion

Our results show that, in all three cases, if the number of travelers between the countries is greater than a certain threshold, the governments switch to a non-restrictive quarantine sooner compared to the socially optimal solution (Figs. 2,3). Namely, in Nash equilibrium, the governors administer the restrictive quarantine for shorter periods compared to the socially optimal solution. In turn, this results in shorter periods before the infection level approaches its full capacity and the restrictive quarantine is administered again (Figs. 2,3). Consequently, under decentralized governance, the solution is sub-optimal, the total amount of time during which a restrictive quarantine is administered is greater, and the average infection level is higher. This result is consistent with previous results that suggested that agents under-invest in the control of diseases [18-23]. Note that in both the optimal solution and the Nash equilibrium, the governments’ actions tend to be synchronized as the second country to switch does so when its infection level is similar to that of the other country (Fig. 2D,F).

**Figure 2:**
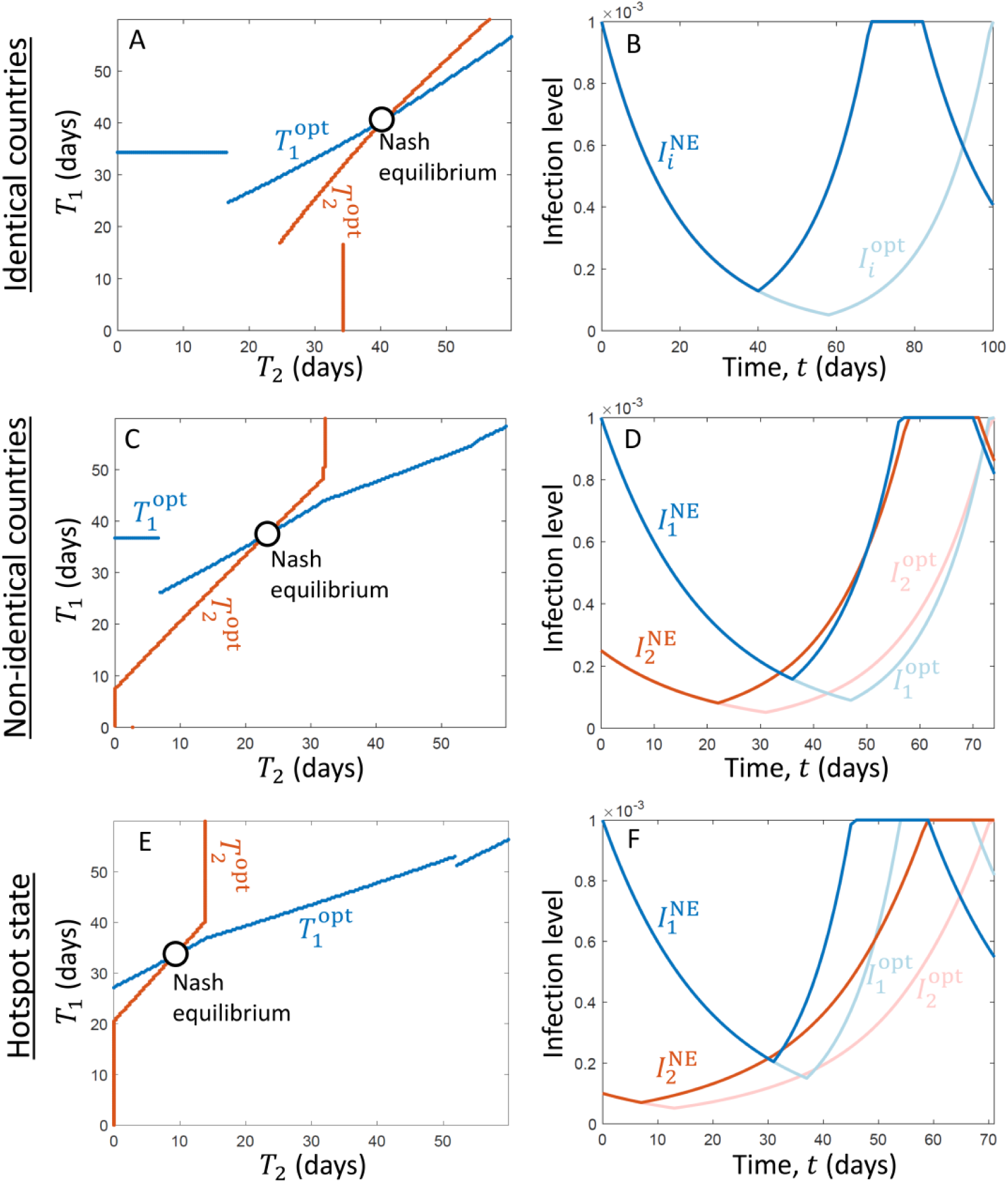
The equilibrium solution dictates that governments release the quarantines earlier than optimal. We consider the three cases illustrated in Fig. 1: identical countries (A, B), non-identical countries (C, D), and a state that initially has a much higher infection level than the rest of the U.S. (E, F). The left column (A, C, E) shows the optimal time for country 1 to release the quarantine, 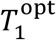, as a function of the day when country 2 releases the quarantine, *T*_2_ (blue line).It also shows the optimal time for country 2 to release the quarantine,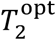, as a function of the day when country 1 releases the quarantine, *T*_1_, on a flipped axis (mirror image, red line). The intersection of the blue and the red line indicates the open-loop Nash equilibrium. In turn, the right column (B, D, E) shows the time evolution of the infection level in countries 1 and 2, assuming that they adopt the Nash equilibrium strategies (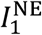, blue line, and 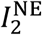, red line), as well as the infection levels assuming that the countries adopt the socially optimal solution (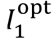, light blue line, and 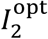, light red line). In all three cases, the governments release the quarantine sooner than the optimum if they follow the Nash equilibrium. Moreover, in both the optimal solution and the Nash equilibrium, the second country to release the quarantine does so approximately when its infection level approaches that of the other country.

**Figure 3:**
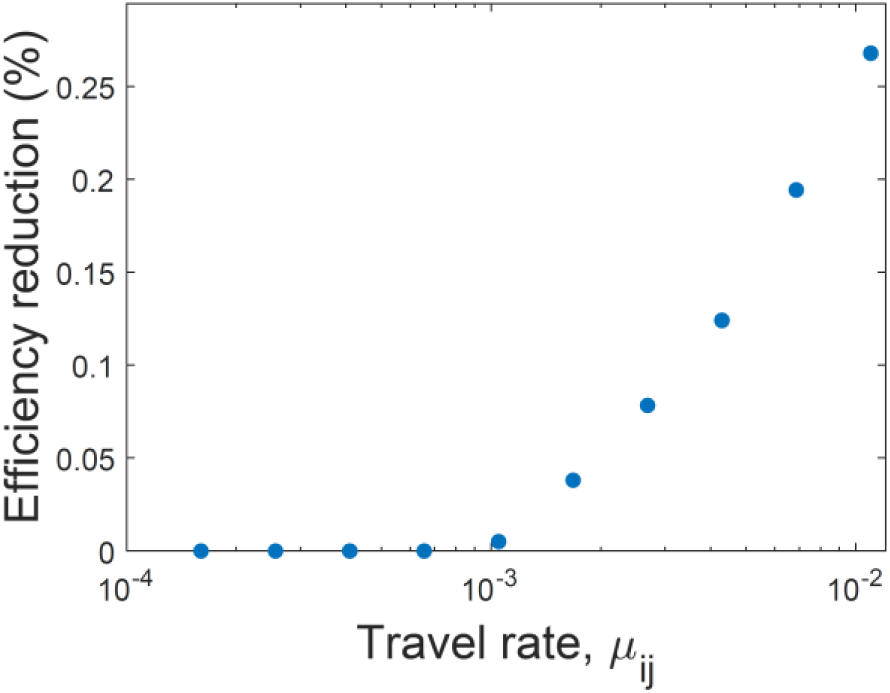
Travel between states may lead to inefficient quarantine policy within each state. Demonstrated is the increase in the relative time that a restrictive quarantine is administered (efficiency reduction, a.k.a. “cost of anarchy”) in a Nash equilibrium (decentralized government) compared to the optimal solution. The efficiency reduction is zero if the proportion of travelers in the population (travel rate, *μ*_*ij*_) is below a certain threshold, and increases with *μ*_*ij*_ above that threshold.

The difference between the optimal solution and the Nash equilibrium emerges if the number of travelers between the countries is above a certain threshold, in which case the inefficiency (price of anarchy) increases with the number of travelers (Fig. 3). However, if the number of travelers is below the threshold, the Nash equilibrium and the optimal solution are identical (Fig. 3). This suggests that one way to prevent the inefficiency due to the decentralized governance is to restrict international or interstate travel to a low level even when the quarantines are non-restrictive.

There are two mutually dependent reasons why decentralized governance results in releasing the restrictive quarantine sooner. First, each governor ignores the damages that its own travelers inflict on other countries, and thus, keeping a higher level of infection is perceived by the governor as less costly. Second, as a consequence of the first reason, each country hosts more infected travelers from the other country, and its own travelers are also hosted in countries with higher infection levels. Consequently, if the infection level in a given country is low, it increases rapidly due to travel, and therefore, it is not worthwhile for the country to reduce its infection level beyond a certain value (where due to travel, this value may be lower than *I*_min_).

Finally, note that we have made numerous simplifying assumptions in our model, which suggests various future directions for examining the consequences of relaxing these assumptions. First, we considered only two types of quarantine, whereas in reality, more options are available. In particular, governments can try to administer an intermediate level of quarantine that keeps the infection at a constant level. Examining whether this policy is better than the ones that we considered is beyond the scope of this paper (see [2, 3, 29]); however, a similar result will likely hold: Decentralized governance might maintain a higher infection level than the optimum. Second, we assumed that travel is allowed under non-restrictive quarantine. However, it would be interesting to examine policies that also dictate how to best integrate the quarantine policy with travel policy. Specifically, further restrictions on travel might mitigate the problem (Fig. 3); however, restrictions on travel come with economic costs [13]. Third, we considered only two countries, whereas considering more countries is generally expected to increase the price of anarchy [30]. And fourth, we considered open-loop solutions in which the governments pre-determine their policy, but communication among the governments might lead to the formation of agreements and coordination of a more global quarantine policy.

## Data Availability

Some parameter values are taken from estimates in the literature and some are estimated from publically available datasets (see Method: Parameterization). All the data and parameter values needed to regenerate the results shown in the figures are given in Methods.

## Supplementary Online Materials

### Methods

#### Dynamics of the infection levels

Here we describe in detail the dynamics of *I*_1_(*t*) and *I*_2_(*t*), which characterize the proportions of infected individuals in the populations of countries 1 and 2, respectively. Assume that each of the two countries can be under a restrictive quarantine or a non-restrictive quarantine, and the type of quarantine can change over time. The dynamics of *I*_*i*_ at time *t* depend on the type of quarantine administered in both countries at that time.

##### <u>Option 1: Country</u> *<u>i</u>*<u>is under a restrictive quarantine</u>

If country *i* is under a restrictive quarantine at time *t, I*_*i*_(*t*) declines at a daily rate −*r*_0_, regardless of what happens in the other country (no travel). However, there are two exceptions. First, *I*_*i*_ cannot decline below a certain threshold, *I*_min_, characterizing the minimal level of infection in the population. Second, if the government switches from restrictive to non-restrictive quarantine, *I*_*i*_ starts to decline only after a delay of *T*_delay_ days. Therefore, in summary,

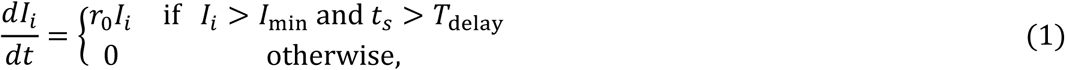

where *t*_*s*_ is the time (in days) from the day when the quarantine became restrictive.

##### <u>Option 2: Country</u> *<u>i</u>*<u>is under a non-restrictive quarantine, but the other country,</u> *<u>j</u>*<u>≠</u> *<u>i</u>*<u>, is under a restrictive quarantine</u>

In this case, there is still no travel between the countries due to the restrictive quarantine in country *j*. Consequently, *I*_*i*_ grows exponentially at a daily rate *r*_*i*_:

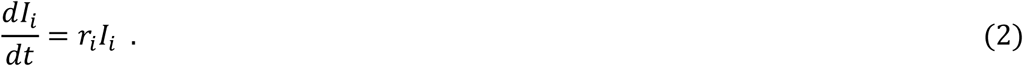

##### Option 3: Both countries are under a non-restrictive quarantine

In this case, individuals travel between the countries, where *μ*_*ij*_ of the residents of country *i* are in travel to country *j*. If we assume that the number of travelers from a country is much smaller than the number of the country’s own residents (*μ*_*ij*_ ≪ *N*_*i*_ and *μ*_*ji*_ ≪ *N*_*j*_), it follows that

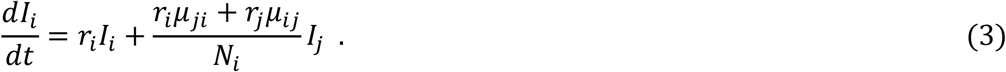

Specifically, (i) the term *r*_*i*_*I*_*i*_ characterizes residents of country *i* that get infected inside country *i* by other residents of country *i*; (ii) the term *r*_*i*_*μ*_*ji*_/*N*_*i*_ characterizes residents of country *i* that get infected inside country *i* by residents of country *j*; and (iii) the term *r*_*j*_*μ*_*ij*_/*N*_*i*_ characterizes residents of country *i* that get infected inside country *j* by residents of country *j*. Note that we assume that travel is temporary (Lagrangian approach [31, 32]); that is, a traveler from country 1 to country 2 is still a resident of country 1 who will return to country 1, and therefore, is s/he gets infected, it increases *I*_1_, even the infection has occurred in country 2.

In turn, without the simplifying assumption that *μ*_*ij*_ ≪ *N*_*i*_, Eq. (3) can be written in a more general form:

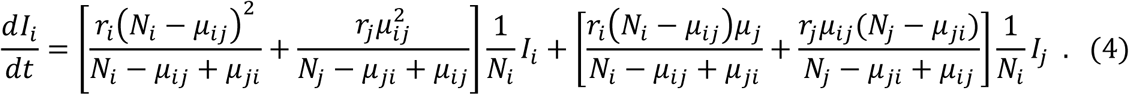

In our simulations, we used values of *μ*_*ij*_ that are much smaller than *N*_*i*_ (see Parameterization subsection), and therefore, the results obtained using Eq. (3) and those obtained using Eq. (4) were almost indistinguishable.

#### Objective functions

The government in each country chooses the time periods during which each type of quarantine is administered in that country. We assume that the objective of each government is to maximize the relative time during which the quarantine is non-restrictive in its country, under the constraint that the health system does not collapse, and therefore, a restrictive quarantine has to be administered in country *i* whenever *I*_*i*_ approaches *I*_max_ [29].

Specifically, denote 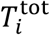 as the time to complete a cycle during which *I*_*i*_ increases to *I*_max_ and returns back to its initial value, *I*_*i*_(0). In turn, denote 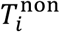 as the total time within such a cycle during which a non-restrictive quarantine is administered in country *i*. Then, we define the utility of country *i* as

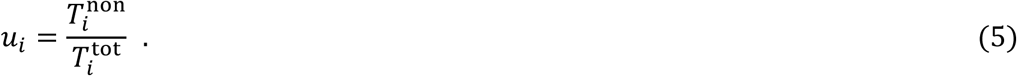

In turn, we calculate both the optimal solution and the open-loop Nash equilibrium. Specifically, the optimal strategy is the one that maximizes *N*_1_*u*_1_ + *N*_2_*u*_2_. The open-loop Nash equilibrium is given by the set of pre-determined strategies that are such that no country can increase its own utility by unilaterally changing its strategy.

#### Numerical methods

Since the growth rates of *I*_1_ and *I*_2_ are piecewise-linear, and since we considered a time delay after the government switches to restrictive quarantine before the infection level starts to decline, it follows from Pontryagin’s maximum principle [30, 33] that the optimal strategy of each government is to choose a time, *T*_*i*_ ≥ 0, at which it switches to a restrictive quarantine and then to wait until *I*_*i*_ = *I*_max_ before switching to a restrictive quarantine again (Fig. 2).

Then, 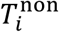, is the period from *T*_*i*_ until *I*_*i*_ approaches *I*_max_, and the time during which the quarantine is restrictive, 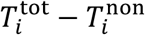, is given by the sum of three distinct periods: (i) between *t* = 0 and *t* = *T*_*i*_, (ii) *T*_*delay*_ days after *I*_*i*_ approaches *I*_max_, and (iii) the time until *I*_*i*_ declines back to its initial value, *I*_*i*_(0).

To find the optimal solution and the open-loop Nash equilibrium, we first generate a matrix in which each cell characterizes a set of strategies, (*T*_1_, *T*_2_), and we simulate dynamics to calculate the utility for each country in every cell. Then, the optimal solution is given by *T*_1_ and *T*_2_ that correspond to the cell in which *N*_1_*u*_1_ + *N*_2_*u*_2_ is maximized. In turn, the open-loop Nash equilibrium is found by calculating the best response of each country to every strategy of the other country, where the intersection of the curves characterizes the Nash equilibrium (Fig. 2 A,C,E). The values of *T*_1_ and *T*_2_ in the matrix vary between 0 and *T*_max_, where *T*_max_ is the time when a restrictive quarantine reduces the infection level in both countries to *I*_min_. (It is never worthwhile to keep a restrictive quarantine for more than *T*_max_ days.) In turn, the size of the matrix determines the resolution at which the strategies *T*_1_ are *T*_2_ are examined, and in Fig. 2, we used a 200 × 200 matrix.

#### Parameterization

The estimations of the parameter values used for the simulation are taken from datasets and literature related to outbreaks of COVID-19 during quarantines [4, 14, 16, 24-27]. Notice that some parameter values vary significantly between countries and states, and therefore, we performed sensitivity analyses to verify that the main results are general and hold within wider parameter ranges.

Under restrictive quarantine conditions, a daily decline rate of 5% in the infection level after the first two weeks is a reasonable estimate. For example, in China, the number of infected individuals declined from ∼86,000 to ∼3,000 within 76 days, which implies *r*_0_ ≈ −5% per day. In turn, in several countries, it took about 14-21 days before any decline occurred following a quarantine, which suggests that considering *T*_*delay*_ ≈ 14 days is reasonable. Next, note that *r*_*i*_ depends on the restrictions used in a given non-restrictive quarantine, and it may vary between states and countries. We used estimates that reflect the weeks before a restrictive quarantine was administered in countries in Europe, where 5% ≤ *r*_*i*_ ≤ 15% per day is a reasonable estimate.

In turn, we assume that *I*_max_ is given approximately by the infection level that was approached in countries in Europe before the restrictive quarantine was administered or two weeks after it was administered, where considering *I*_min_ ≈ 0.01% − 0.1% of the total population size is a reasonable estimate. Next, parameters like *I*_min_ and the travel rates *μ*_12_ and *μ*_21_ are harder to estimate, as they depend on the specific location and scale of the countries and states considered. Therefore, we performed sensitivity analyses and examined a variety of values (e.g., Fig. 3). Finally, the ratio between *N*_1_ and *N*_2_ reflects the relative population sizes of the two countries: *N*_1_ = *N*_2_ characterizes the cases demonstrated in Fig. 1A,B and 2A-D, while *N*_1_ < *N*_2_ characterizes the case demonstrated in Fig. 1C and 2E-F.

#### Parameter values used to generate the figures

<u>Fig. 2A,B:</u> *r*_0_ = −5% (day_^−1^_), *r*_1_ = *r*_2_ = 7% (day_^−1^_), *I*_max_ = 0.1%, *I*_min_ = 0.005%, *T*_*delay*_ = 14 (days), *N*_1_ = *N*_2_ = 1, *μ*_12_ = *μ*_21_ = 0.2% × *N*_1_.

<u>Fig. 2C,D:</u> *r*_0_ = −5% (day_^−1^_), *r*_1_ = 9% (day_^−1^_), *r*_2_ = 7% (day_^−1^_), *I*_max_ = 0.1%, *I*_min_ = 0.005%, *T*_*delay*_ = 14 (days), *N*_1_ = *N*_2_ = 1, *μ*_12_ = *μ*_21_ = 0.25% × *N*_1_.

<u>Fig. 2E,F:</u> *r*_0_ = −5% (day_^−1^_), *r*_1_ = 11% (day_^−1^_), *r*_2_ = 5% (day_^−1^_), *I*_max_ = 0.1%, *I*_min_ = 0.005%, *T*_*delay*_ = 14 (days), *N*_1_ = 0.3, *N*_2_ = 1, *μ*_12_ = 0.25% × *N*_1_, *μ*_21_ = 0.25% × *N*_2_.

<u>Fig. 3:</u> *μ*_12_ = *μ*_21_ varies (x-axis), and the other parameters are the same as those in Fig. 2A,B.

